# Accuracy of pattern-based dementia diagnostic protocols: Using longitudinal data to infer etiology of Alzheimer’s disease and related dementia as compared to stroke or normal aging

**DOI:** 10.1101/2025.02.26.25322969

**Authors:** Sean A. P. Clouston, Douglas W. Hanes, Mahdieh Danesh Yazdi

**Affiliations:** Program in Public Health and Department of Family, Population, and Preventive Medicine, Renaissance School of Medicine at Stony Brook University, Stony Brook, NY, USA

## Abstract

**Introduction:** The goal of this study was to compare accuracy for pattern-recognition protocols to prospectively identify dementia due to Alzheimer’s disease and related dementias (ADRD) or stroke.

**Methods:** We used data from cognitively unimpaired respondents who completed at least 5 cognitive assessments during waves 3–12 of the *Health and Retirement Study* (HRS), a longitudinal study of older US residents. Participants were assessed at wave 12 (in 2019) for cognitive status. Patterns of participants’ cognitive decline were analyzed to differentially identify ADRD and stroke and were compared against self-reported and objective diagnoses of amnestic, executive, and multidomain mild cognitive impairment (MCI) and ADRD. We reported sensitivity/specificity to detect new-onset dementia at the final wave of observation.

**Results:** After applying inclusion/exclusion criteria, 43 (1.69%) of cognitively unimpaired participants developed dementia, while 165 (6.49%) developed amnestic MCI. Patterns of cognitive decline consistent with ADRD affected 372 (14.6%) of respondents, while patterns of cognitive decline consistent with stroke were evident in 917 (36.1%) participants. ADRD- consistent cognitive declines were evident in 75.8% and 76.7% cases of amnestic MCI and dementia, respectively, though only 24.5% reported receiving a clinical diagnosis of dementia. Sensitivity/Specificity of ADRD was 94.3%/87.0% when detecting dementia without stroke.

**Discussion:** This study implies that we can reliably use longitudinal patterns of cognitive decline to differentially diagnose ADRD from Stroke in most participants with dementia.

## Background

Alzheimer’s disease and related dementias (ADRD) was the seventh-most common cause of death in 2022, while stroke was fifth ^1^. Both stroke and ADRD cause both mild cognitive impairment (MCI) and dementia ^2^, so attributing etiology to the disease can be difficult.

Neurocognitive differentiation is possible using memory loss to indicate the presence of ADRD and executive dysfunction to identify stroke; however, both stroke and ADRD can cause cognitive decline across both domains alongside changes in orientation and language, making definitive diagnosis difficult when relying on these tools alone.

Stroke and ADRD are clinically differentiated based on a symptom history that seeks to determine the temporal relationship with cognitive performance (Figure 1).^3^ Whereas ADRD causes a progressively accelerated rates of cognitive decline,^4^ stroke causes a rapid loss in cognition that may be sustained but does not worsen progressively.^5^ Seeking to use the characteristics of longitudinal cognitive trajectories to aid in differential diagnosis, recent work has proposed using pattern recognition programs to help identify individuals suffering from cognitive declines consistent with these diseases.^3^ This work reports that these programs can predict self-reported diagnosis of dementia ^6^ or stroke ^7^ and accounts for approximately half of all cognitive aging.^3^ To date, little is known about the sensitivity/specificity of these protocols. The goal of this study was to compare the sensitivity and specificity of different diagnostic protocols to prospectively identify different causes of dementia. Secondarily, we sought to examine the long-term prognosis of the diagnostic protocol.

**Figure 1.**
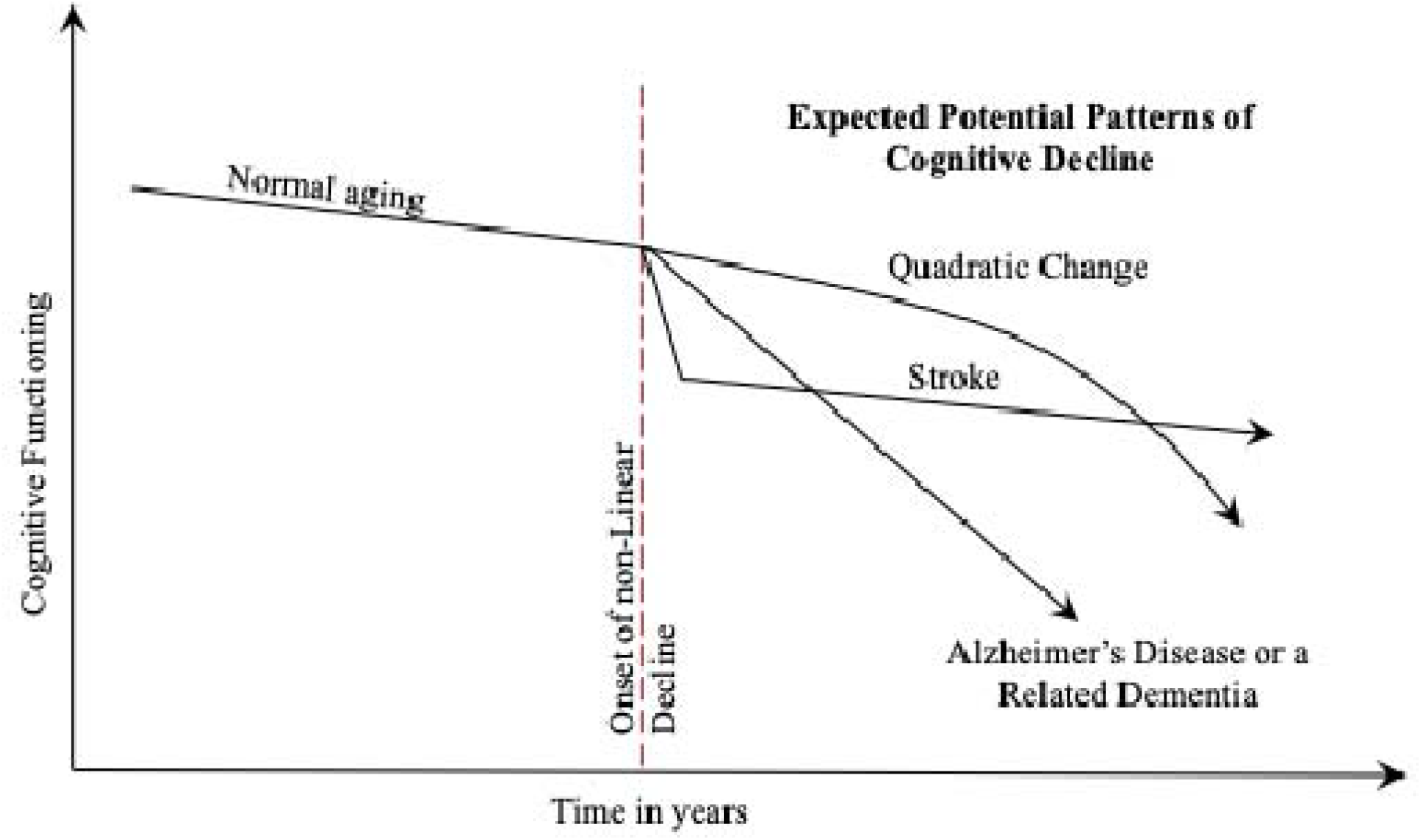
Cognitive profiles of different pathological processes.

## Methods

In this study, we used data from the *Health and Retirement Study* (HRS), a longitudinal study of older US residents (response rate 81.6%) that includes biannual, multi-domain objective cognitive assessments as well as self-reported measures of cognitive health. We used waves 3-12 that are publicly available online and exclude data from waves occurring near to COVID-19 (http://hrsonline.isr.umich.edu) ^8^, because cognitive measures were stable throughout this period. Self-reported dementia diagnosis was only available for waves 10-12, so ultimate outcomes were defined at wave 12 though prior self-reported diagnosis may also be reported in earlier waves among individuals who attrite.

### Inclusion/exclusion

To successfully assess the pattern of cognitive decline we included individuals who completed ≥5 cognitive performance assessments. To ensure that dementia diagnosis was feasible, we excluded individuals with evidence of possible MCI or dementia at baseline and those who completed the full battery of tasks prior to their last survey. Finally, since the neuropathological cascade in Alzheimer’s disease takes 7-15 years to develop into dementia ^9^, we focused this analysis on those individuals whose diagnoses of stroke occurred prior to their final observations and those individuals whose onset of accelerated cognitive decline occurred >6 years prior to diagnosis of dementia.

## Measures

Cognitive functioning measures in the HRS include measures of memory and executive functioning that are most susceptible to cognitive pathology ^10^ and are measured using a modified version of the telephone interview for cognitive status ^11,12^. *Episodic memory* is the summation of verbal learning and recall tasks. Verbal learning asks individuals to learn a list of ten words and repeat as many back to the interviewer correctly as possible (/10). Verbal recall asks respondents to again recall the list of words around 12 minutes later, following intermediary questioning (/10). *Executive Function* was measured as the summation of the serial 7s subtraction test (/5), the backwards counting task (/2). Functional limitations were calculated using disorientation measures including the name of current President/Vice-President (/2), names of two common objects provided verbal descriptions (/2), and the correct date (day, month, year) and day of the week (/4).

*Diagnostic Outcomes* in this study include potential self-reported diagnosis of ADRD or stroke. Participants are often lost to follow-up due to poor cognitive status ^13^, so in order to substantially reduce attrition bias, we stratified outcomes by whether responses were completed by proxy respondents ^14^. Among individuals who did not report a diagnosis we identified whether they left the study before the next follow-up or if their next follow-up was completed by a proxy respondent. Finally, following NIA-AA guidelines we defined MCI as the presence of evidence of cognitive impairment (<1.5 SD of average functioning) in amnestic or executive domains ^15^. We also identified a multidomain category by the presence of impairments across both cognitive domains. To identify dementia, we categorized outcomes by the presence of multi-domain cognitive impairment and the presence of functional limitations. Because participants’ MCI/dementia status was not available at the time of the proxy response, we carried forward the most recent cognitive-based diagnosis to the final wave, where necessary making it possible for a person to both have a positive diagnosis of dementia and to have a proxy respondent.

*Patterns of Cognitive Decline* were indicated using maximum likelihood estimation on episodic memory trajectories. Specifically, for everyone with ≥5 cognitive observations, we identified the best-fitting potential change-point for every respondent. However, because it is unlikely that all respondents will experience ADRD, we also fit other potential patterns including: the “step pattern” commonly reported in stroke, a linear pattern consistent with normal aging, and a quadratic model consistent with underlying theory implied by quadratic random slopes models. Akaike’s information criteria (AIC) was used to compare pathological and linear models. In a small number of cases, linear patterns were observed, but the rate of change was more consistent with accelerated onset in ADRD than with normal aging. Since these slopes were unsustainable, we decided to use an optimal cut-point of -0.16 to characterize these as post-acceleration ADRD.

### Statistical Analysis

We began by providing means and standard deviations or frequencies and percentages to describe the sample characteristics. Next, we stratified by most recent cognitive outcome in participants and for each pattern of cognitive decline we calculated sensitivity, specificity, and positive predictive value to identify the presence of dementia in the last observed year or at wave 12. To examine the range of potential self-reported outcomes, we then stratified by self-reported diagnoses and reported the percentages of cases that were explained by each pattern of cognitive decline.

### Ethics

The institutional review board determined that this secondary data analysis of publicly accessible data did not constitute human subjects’ research.

## Results

We began by applying inclusion/exclusion criteria, resulting in an analytic sample of 2,543 individuals. Examining sample characteristics for this study and found that individuals in the study were in their early fifties at baseline. The average person had more than a high school education, and most participants reported White race/ethnicity.

We examined incident cognitive status in participants without any evidence of cognitive impairment at baseline stratified by cognitive diagnosis (Table 2). Stroke was the most common outcome, and, among strokes, the most common outcome was a lack of cognitive impairment. In contrast, most individuals with cognitive profiles consistent with ADRD had evidence of cognitive impairment most often with amnestic presentation. Results for this study revealed that ADRD-like accelerated cognitive declines accounted for 75.8% of cases of amnestic MCI and 76.7% of cases of dementia. Evidence of stroke in the cognitive profile accounted for 40.0% of cases of executive MCI and 18.6% of cases of dementia. The ADRD diagnostic routines had excellent sensitivity, specificity, and positive predictive value for detecting a person who would go on to develop dementia. In contrast, quadratic and linear forms of cognitive decline both had lower overall sensitivity and explained fewer cases of dementia.

**Table 1.** Sample Characteristics for Health and Retirement Study Participants, n = 2,543.

| Sample Characteristics | Mean (SD) / N<br>(%) |
| --- | --- |
| Age at first assessment, years | 53.87 (5.41) |
| Educational Attainment |  |
| Less than High School | 241 (9.48%) |
| High School Diploma | 963 (37.87%) |
| Some College | 645 (25.36%) |
| University Degree | 694 (27.29%) |
| Female | 866 (34.05%) |
| Race/Ethnicity |  |
| White | 2,148 (84.47%) |
| Black | 215 (8.45%) |
| Hispanic | 46 (1.81%) |
| Other | 134 (5.27%) |

**Note:** SD: Standard deviation; %: percentage; N: frequency

**Table 2.** Diagnostic outcomes among individuals who completed dementia assessment at final wave.

|  | Pattern Recognition Identification |  |  |  |
| --- | --- | --- | --- | --- |
|  | None<br>(n=1113<br>[43.77%]) | Quadratic<br>(n=141<br>[5.54%]) | Stroke<br>(n=917<br>[36.06%]) | ADRD<br>(n=372<br>[14.63%]) |
| Cognitively Unimpaired | 948<br>(49.09%) | 119<br>(6.16%) | 722<br>(37.39%) | 142<br>(7.35%) |
| Amnesic MCI | 5<br>(3.03%) | 2<br>(1.21%) | 33<br>(20.00%) | 125<br>(75.76%) |
| Executive MCI | 157<br>(47.15%) | 20<br>(6.01%) | 133<br>(39.94%) | 23<br>(6.91%) |
| Multidomain MCI | 1 (1.41%) | 0 (0%) | 21<br>(29.58%) | 49<br>(69.01%) |
| Dementia | 2 (4.65%) | 0 (0%) | 8<br>(18.60%) | 33<br>(76.74%) |
| <u>Dementia</u> |  |  |  |  |
| Accuracy |  | 88.7% | 56.9% | 87.2% |
| Sensitivity |  | 0.0% | 80.0% | 94.3% |
| Specificity |  | 88.8% | 56.8% | 87.0% |

**Note:** %: percentage; N: frequency

Stratification of results by self-reported diagnostic status (Table 3) revealed that 17.7% of cases of ADRD-like cognitive decline with evidence of prevalent dementia reported receiving a diagnosis. This rate increased to 41.7% among individuals who had cognitive decline consistent with a stroke. Intriguingly, 15.9% of those whose cognitive profile was consistent with a stroke and who had evidence of executive MCI, multidomain MCI, or dementia reported a diagnosis of stroke. Most cases with evidence of ADRD or stroke in their cognitive profile reported no neurological diagnosis.

**Table 3.** Diagnostic outcomes stratified by self-reported diagnosis among individuals who completed dementia assessment at final wave.

|  | Pattern Recognition Identification |  |  |  |
| --- | --- | --- | --- | --- |
|  | None | Quadratic | Stroke | ADRD |
| No Reported Diagnosis | N (%) | N (%) | N (%) | N (%) |
| Cognitively Unimpaired | 890 (50.54%) | 110 (6.25%) | 638 (36.23%) | 123 (6.98%) |
| Amnesic MCI | 5 (3.94%) | 1 (0.79%) | 30 (23.62%) | 91 (71.65%) |
| Executive MCI | 143 (48.31%) | 20 (6.76%) | 112 (37.84%) | 21 (7.09%) |
| Multidomain MCI | 1 (1.85%) | 0 (0%) | 15 (27.78%) | 38 (70.37%) |
| Dementia | 1 (4%) | 0 (0%) | 4 (16%) | 20 (80%) |
| Patient-Reported Dementia Diagnosis |  |  |  |  |
| Cognitively Unimpaired | 2 (9.09%) | 6 (27.27%) | 3 (13.64%) | 11 (50%) |
| Amnesic MCI | 0 (0%) | 1 (4.55%) | 10 (45.45%) | 11 (50%) |
| Executive MCI | 1 (12.5%) | 2 (25%) | 1 (12.5%) | 4 (50%) |
| Multidomain MCI | 0 (0%) | 1 (12.5%) | 3 (37.5%) | 4 (50%) |
| Dementia | 0 (0%) | 0 (0%) | 3 (50%) | 3 (50%) |
| Patient-Reported Stroke |  |  |  |  |
| Cognitively Unimpaired | 56 (35.44%) | 9 (5.7%) | 77 (48.73%) | 16 (10.13%) |
| Amnesic MCI | 0 (0%) | 1 (4%) | 2 (8%) | 22 (88%) |
| Executive MCI | 13 (40.63%) | 0 (0%) | 18 (56.25%) | 1 (3.13%) |
| Multidomain MCI | 0 (0%) | 0 (0%) | 3 (27.27%) | 8 (72.73%) |
| Dementia | 1 (8.33%) | 0 (0%) | 3 (25%) | 8 (66.67%) |
| Patient-Reported Stroke and Dementia |  |  |  |  |
| Cognitively Unimpaired | 1 (50%) | 0 (0%) | 0 (0%) | 1 (50%) |
| Amnesic MCI | 0 (0%) | 0 (0%) | 2 (50%) | 2 (50%) |
| Executive MCI | 1 (50%) | 0 (0%) | 0 (0%) | 1 (50%) |
| Multidomain MCI | 2 (50%) | 0 (0%) | 0 (0%) | 2 (50%) |
| Dementia | 1 (16.67%) | 0 (0%) | 2 (33.33%) | 3 (50%) |
| Diagnosis Rate | 33.33% | – | 41.67% | 17.65% |

**Note:** %: percentage; N: frequency. Individuals who were lost to follow-up prior to diagnosis were recorded as having no reported diagnosis.

## Discussion

The goal of this study was to examine what happened to participants who were identified with a pattern-recognition program as having a cognitive profile consistent with stroke or ADRD. Our main interest was in assessing accuracy to detect objective and self-reported diagnostic status in the decade following onset. Our study found that accuracy was high for the ADRD pattern-recognition protocol, with overall accuracy, sensitivity, and specificity exceeding 85%. Similarly, we found that individuals identified as having had a stroke were also common, but accuracy of detecting dementia was reduced in those with stroke. Overall, these two patterns of cognitive decline accounted for 95.3% of all cases of dementia identified in the study and 91.7% of all cases of dementia that were corroborated by self-reported dementia diagnosis. Together, these results suggest that cognitive profiling has a high degree of accuracy to predict the onset of dementia.

While most participants with an ADRD-like pattern of cognitive decline had evidence of MCI or dementia, a relatively large number of individuals with ADRD-like patterns of cognitive decline did not experience onset of dementia after 10 years, with many remaining cognitively unimpaired while others had only earned diagnoses of amnestic or multidomain MCI. While true, it is worth noting that 65.6% of all cases identified as ADRD had converted to a clinically significant diagnostic status, most with an amnestic phenotype. These results corroborate a large literature proposing the utility of amnestic MCI as a prodrome of ADRD.^16^ Results also suggest that, even though there is evidence of ADRD-like acceleration in the cognitive profile, this is not necessarily going to lead to dementia within the short timeframe shown here. This suggests the potential for more muted prognoses in up to one-third of all cases.

This study further corroborates evidence suggesting that executive MCI may be a phenotype particularly damaged by stroke. Yet our results also revealed that many individuals with cognitive profiles consistent with stroke also received diagnoses of amnestic MCI or, more commonly, multidomain MCI. These results imply that amnestic MCI can, in a minority of cases, be a consequence of multiple different etiologies and, therefore, could have variable predictive power for dementia. Indeed, in our study we examined the predictive power of amnestic MCI for dementia and found that the sensitivity/specificity for amnestic MCI to detect dementia was much higher in individuals with ADRD-like declines but more moderate in those lacking ADRD-like cognitive declines. As a result, the pattern identification was usually more specific but was also always more accurate for detecting dementia than amnestic MCI.

Our results support prior studies suggesting widespread under-diagnosis of both stroke and ADRD.^17^ In the present study, we determined that only 17.7% of cases of accelerated cognitive decline that resulted in widespread cognitive impairment consistent with ADRD reported a diagnosis of stroke or dementia. Slightly better, 41.7% of cases of stroke were diagnosed as stroke or dementia. Similarly, we found that only 37% of cases of stroke comorbid with executive MCI or more severe levels of cognitive impairment were ultimately diagnosed. Diagnosis reporting rates are low, suggesting the potential that research relying solely on medical diagnoses may be subject to severe observational biases in cases where diagnosis is not systematic.

### Limitations

While seeking to understand longitudinal cognitive profiles in one of the largest and longest-running cognitive monitoring studies in the world, this study is limited in several ways. First, to attempt cognitive profiling, this study needed five cognitive waves that are assumed to begin before the onset of either condition. However, it is impossible to determine with any degree of certainty if disease onset occurred at the time observed or before the observational window. This type of observational window bias may be problematic and could result in missed diagnoses. Notably, several hundred individuals identified as having no evidence of any stroke had linear rates of decline resulting in evidence of executive MCI. Interestingly, of those with linear rates of decline who reported a stroke, the majority had executive MCI at follow-up. These results may imply either that many strokes predate data-collection or, in contrast, that executive MCI is common in the general population. Second, while this study sought to use diagnostic routines to mimic medical diagnoses, there is no way to exclude conditions like chronic traumatic encephalopathy, brain cancer, alcohol-related dementia, or psychosis that might cause cognitive decline in a functionally similar way. We note that these conditions tend to be rare and are therefore unlikely to be a major source of bias in this study. Third, in examining self-reported diagnosis we are open to the potential for recall bias especially given that we are studying a group of individuals with cognitive impairment. Underreporting of these diagnoses would result in under-estimates of the risk of diagnosis. Since these rates are so low, we do not believe that this recall bias would substantially change the result of this study.

### Impact

Better identifying dementia, earlier, has been a longstanding goal in the research community. In this study, we went further to determine to what extent we could use cognitive data alone to differentially diagnose stroke versus ADRD and, if so, to report the accuracy of diagnosis using both objective and patient-reported outcomes. Our results implied that longitudinal data can be used to begin to differentiate the etiology of cognitive decline in a reliable and non-invasive way in cases where substantial longitudinal data are available.

Conveniently, this is becoming more common since large care providers like Medicare have started to recognize the importance of cognitive monitoring for older populations. Clinical work might therefore benefit from monitoring not only cognitive function but also the rate and timing of cognitive decline to better identify cases earlier and in an objective and comprehensive manner.

## Data Availability

All data in the present study are available online at hrs.isr.umich.edu

https://hrs.isr.umich.edu/data-products

